# Clinical characteristics and outcomes in diabetes patients admitted with COVID-19 in Dubai: a cross-sectional single centre study

**DOI:** 10.1101/2020.07.08.20149096

**Authors:** R Bhatti, HK Amar, S Khattib, S Shiraz, G Matfin

## Abstract

**Aim:** To describe the clinical characteristics and outcomes of hospitalised Coronavirus Disease 2019 (COVID-19) patients with diabetes.

**Methods:** A cross-sectional observational study was conducted in patients with diabetes admitted with COVID-19 to Mediclinic Parkview Hospital in Dubai, United Arab Emirates (UAE) from 30^th^ March to 7^th^ June 2020. They had laboratory and/or radiologically confirmed severe acute respiratory syndrome-coronavirus-2 (SARS-CoV-2), known as COVID-19. Variation in characteristics, length of stay in hospital, diabetes status, comorbidities and outcomes were examined.

**Results:** A total of 103 patients with confirmed COVID-19 presentations had diabetes. During the same timeframe, 410 patients overall were admitted with COVID-19 infection. This gives a total proportion of persons admitted with COVID-19 infection and coexistent diabetes/prediabetes of 25%. 67% (n=69) of the COVID-19 diabetes cohort were male. Patients admitted with COVID-19 and diabetes represented 17 different ethnicities. Of these, 59.2% (n=61) were Asians and 35% (n=36) were from Arab countries. Mean age (SD) was 54 (±12.5) years. 85.4% (n=88) were known to have diabetes prior to admission, while 14.6% (n=15) were newly diagnosed with either diabetes or prediabetes during admission. Most patients in the study cohort had type 2 diabetes or prediabetes, with only 3% overall having type 1 diabetes (n=3). 46.9% of patients had evidence of good glycaemic control of their diabetes during the preceding 4-12 weeks prior to admission as defined arbitrarily by admission HbA1c <7.5%. 73.8% (n=76) had other comorbidities including hypertension, ischaemic heart disease, and dyslipidaemia. Laboratory data (Mean ± SD) on admission for those who needed ward-based care versus those needing intensive care unit (ICU) care: Fibrinogen 462.75 (±125.16) mg/dl vs 660 (±187.58) mg/dl ; D-dimer 0.66 (±0.55) µg/ml vs 2.3 (±3.48) µg/ml; Ferritin 358.08 (±442.05) mg/dl vs 1762.38 (±2586.38) mg/dl; and CRP 33.9 (±38.62) mg/L vs 137 (±111.72) mg/L were all statistically significantly higher for the ICU cohort (p<0.05). Average length of stay in hospital was 14.55 days. 28.2% of patients needed ICU admission. 4.9% (n=5) overall died during hospitalisation (all in ICU).

**Conclusions:** In this single-centre study in Dubai, 25% of patients admitted with COVID-19 also had diabetes/prediabetes. Most diabetes patients admitted to hospital with COVID-19 disease were males of Asian origin. 14.6% had new diagnosis of diabetes/prediabetes on admission. The majority of patients with diabetes/prediabetes and COVID-19 infection had other important comorbidities (n=76; 73.8%). Only 4 patients had negative COVID-19 RT-PCR but had pathognomonic changes of COVID-19 radiologically. Our comprehensive laboratory analysis revealed distinct abnormal patterns of biomarkers that are associated with poor prognosis: Fibrinogen, D-dimer, Ferritin and CRP levels were all statistically significantly higher (p<0.05) at presentation in patients who subsequently needed ICU care compared with those patients who remained ward-based. 28.2% overall needed ICU admission, out of which 5 patients died. More studies with larger sample sizes are needed to compare data of COVID-19 patients admitted with and without diabetes within the UAE region.

## Introduction

The burden of the Severe Acute Respiratory Syndrome-Coronavirus-2 (SARS-CoV-2), which is known as COVID-19 has being increasing worldwide. According to World Health Organisation (WHO), COVID-19 was declared as an epidemic on 11th March 2020.^1^ To date 49,069 cases have been reported in the United Arab Emirates (UAE) with 316 deaths.^2^

Recent studies have suggested that diabetes is a major risk factor contributing to severity of illness and mortality from COVID-19.^3^ For example, a meta-analysis of seven trials in China showed that 9.7% of 1576 patients with COVID-19 had diabetes.^4^ Diabetes is also a major risk factor for the development of a more severe pneumonia and clinical course due to COVID-19 infection, and occurs in around 20-30% of such patients. ^5^ In addition, poor glycaemic control whether related to diabetes or stress hyperglycemia is also known to be associated with poor patient outcomes including hospitalisation and death. ^6^ In a large population-based study in England, 23,804 COVID-19 related deaths were reported. Of these, one third occurred in people with diabetes: 7,466 (31·4%) with Type 2 and 365 (1·5%) with Type 1 diabetes.^7^

The aim of this Dubai-based, single-centre cross-sectional study was to assess the clinical characteristics and outcomes of patients with diabetes admitted with moderate to severe COVID-19 illness.

## Methods

### Study design

This is a cross-sectional observational study of hospitalised patients with diabetes who had laboratory and/or radiological confirmed COVID-19 disease between 30^th^ March and 7^th^ June 2020 at Mediclinic Parkview Hospital, Dubai, UAE.

### Definitions

A diagnosis of COVID-19 was confirmed with either positive COVID-19 RT-PCR testing and/or consistent imaging findings on Chest X-ray / Chest HRCT scan (i.e. pathognomonic changes of ground glass radiological features of COVID-19).

#### For the purposes of the study

Diabetes was confirmed by either prior diagnosis or admission HbA1c ≥ 6.5%.

Prediabetes was defined as by either prior diagnosis and/or admission HbA1c 5.7-6.4%.

Uncontrolled diabetes was defined as admission HbA1c ≥ 7.5%.

Patients were discharged from hospital when clinically well and two negative laboratory confirmed nasopharyngeal swabs for COVID-19 RT-PCR.

Admission laboratory tests were defined as tests done within 24 hours of hospital admission.

### Data collection

Data was collected from electronic medical records Bayanaty^®^. Information was extracted on basic demographics, nationalities, laboratory data, imaging results (i.e. chest x-ray, chest HRCT), and capillary blood glucose done during admission. Ethical approvals were taken from local Mediclinic Institutional Research Board; and Dubai Scientific Research Ethics Committee, Dubai Health Authority, Dubai, UAE.

### Statistical Analaysis

SPSS (IBM Corp. Released 2019. IBM SPSS Statistics for Windows, Version 25.0. Armonk, NY: IBM Corp) was used for statistical analysis. Frequencies with proportions were reported for categorical variables and means with standard deviations (SDs) were reported for continuous variables. Mann Whitney test is used and p value of less than 0.05 is considered as significant.

## Results

A total of 103 patients admitted to hospital with confirmed COVID-19 had diabetes. During the same timeframe, 410 patients overall were admitted with COVID-19 infection. This gives a total proportion of persons admitted with COVID-19 infection and coexistent diabetes/prediabetes of 25%. 67% (n=69) of the COVID-19 diabetes cohort were male. Patients admitted with COVID-19 and diabetes represented 17 different ethnicities. Of these, 59.2% (n=61) were Asians and 35% (n=36) were from Arab countries. Mean age (SD) was 54 (±12.5) years. 73.8% (n=76) had other comorbidities including hypertension, ischaemic heart disease, dyslipidaemia, and chronic kidney disease (shown in Table 1). 9.7% patients were on ACE-I and 31.1% patients were on ARB on admission, which were continued as per individual clinical circumstances during admission with no obvious adverse effects.

**Table 1:** Basic Demographics and Pre-comorbidities

|  | Items | Total, N (103) |
| --- | --- | --- |
| Demographics | Gender, n (%) |  |
|  | Male | 69 (67) |
|  | Female | 34 (33) |
|  | Nationality, n (%) |  |
|  | Arabic countries | 36 (35) |
|  | Asian countries | 61 (59.2) |
|  | Western countries | 5 (4.9) |
|  | African Countries | 1 (1) |
|  | Age in years, mean (SD) | 54 (12.5) |
| Comorbidities | Comorbidity, n (%) |  |
|  | Yes | 76 (73.8) |
|  | No | 27 (26.2) |
|  | Comorbidities*, n (%) |  |
|  | Hypertension | 66 (64) |
|  | Dyslipidaemia | 54 (52.4) |
|  | Ischemic heart disease | 11 (10.6) |
|  | Breast cancer | 7 (6.8) |
|  | Chronic kidney disease | 4 (3.9) |
|  | Stroke | 3 (2.9) |
|  | COPD | 1 (0.97) |
|  | Renal transplant | 1 (0.97) |
|  | ACE-I or ARB Medications, n (%) |  |
|  | ACE-I | 10 (9.7) |
|  | ARB | 32 (31.1) |

\* More than one disease is possible per patient.

Out of 103 patients, 3 patients (2.9%) had type 1 diabetes and 90 patients (87.4%) had type 2 diabetes. 6/90 patients were newly diagnosed with type 2 diabetes during admission. 10 patients (9.4%) had prediabetes, of which 9 had a new diagnosis on admission. Of those patients with known diabetes diagnosis, 8 patients were on basal insulin regimen and 6 patients on basal bolus insulin regimen (Table 2).

**Table 2:**
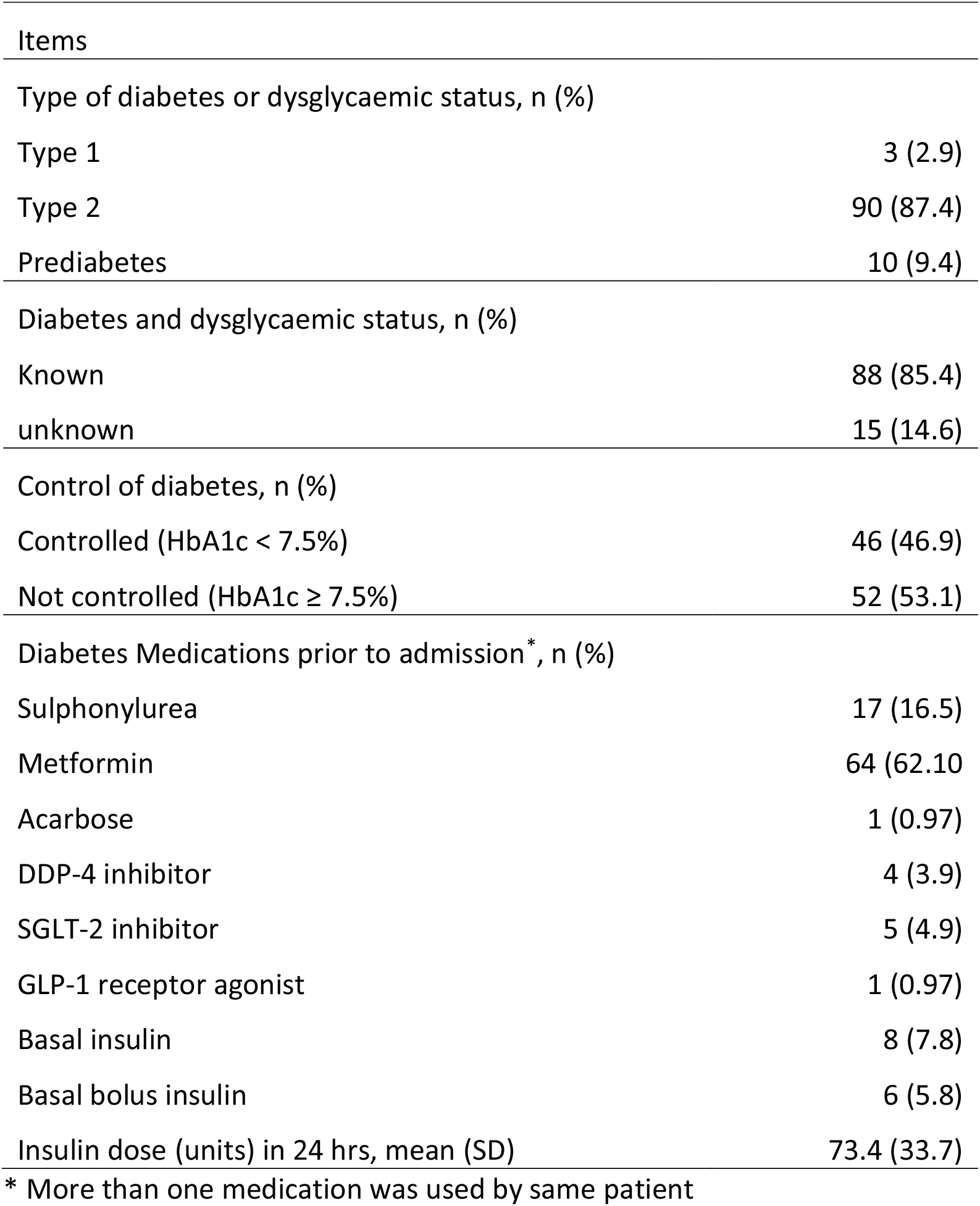
Diabetes manifestation pre-COVID-19 infection

| Items |  |
| --- | --- |
| Type of diabetes or dysglycaemic status, n (%) |  |
| Type 1 | 3 (2.9) |
| Type 2 | 90 (87.4) |
| Prediabetes | 10 (9.4) |
| Diabetes and dysglycaemic status, n (%) |  |
| Known | 88 (85.4) |
| unknown | 15 (14.6) |
| Control of diabetes, n (%) |  |
| Controlled (HbA1c < 7.5%) | 46 (46.9) |
| Not controlled (HbA1c ≥ 7.5%) | 52 (53.1) |
| Diabetes Medications prior to admission*, n (%) |  |
| Sulphonylurea | 17 (16.5) |
| Metformin | 64 (62.10) |
| Acarbose | 1 (0.97) |
| DDP-4 inhibitor | 4 (3.9) |
| SGLT-2 inhibitor | 5 (4.9) |
| GLP-1 receptor agonist | 1 (0.97) |
| Basal insulin | 8 (7.8) |
| Basal bolus insulin | 6 (5.8) |
| Insulin dose (units) in 24 hrs, mean (SD) | 73.4 (33.7) |

\* More than one medication was used by same patient

On admission investigations (Table 3), average random blood glucose in laboratory was 10.2 mmol/L. 56/103 (54%) patients had lymphopenia on admission. 65 patients had confirmed pneumonia radiologically. 38 patients had normal chest x-ray, 14 out of these had confirmed pneumonia on chest HRCT. 4 out of 103 patients were negative on laboratory RT-PCR but had radiologically confirmed COVID-19 pathognomonic changes.

**Table 3:** Laboratory data

| On admission, mean (SD) |  | Normal ranges |
| --- | --- | --- |
| HbA1c % | 7.69 (± 1.56) | <5.6 |
| Random blood glucose (mmol/L) | 10.2 (± 4.2) | 3.89-7.70 |
| Hb (g/dl) | 12.8 (± 1.73) | 13-17.5 |
| White cell count ($10^3/\mu\text{L}$) | 6.5 (± 3.11) | 4-11 |
| Lymphocyte Count ($10^3/\mu\text{L}$) | 1.24 (± 0.76) | 1-4.80 |
| Lymphopenia n (%) | 56 (54.3) |  |
| Platelets ($10^3/\mu\text{L}$) | 240.38 (±85.30) | 150-450 |
| Creatinine ($\mu\text{mol/L}$) | 92.74 (±48.92) | 63.6-110.5 |
| GFR (ml/min/$1.73^2$) | 82.86 (±22.2) | >60 |
| Ferritin (ng/ml) | 757.34 (±1548.88) | 21.81-274.66 |
| Creatine Kinase (U/L) | 402.2(±1087.60) | 30-200 |
| LDH (U/L) | 309.32 (±228.96) | 125-243 |
| Fibrinogen (mg/dl) | 529.29 (±175.23) | 200-400 |
| Procalcitonin | 0.63 (±1.24) | |
| CRP (mg/L) | 64.15 (±82.7) | 0-5 |
| D-dimer ($\mu\text{g/ml}$) | 1.16 (±2.09) | 0-0.5 |

Emergent treatments used to manage diabetes patients with COVID-19 and related outcomes are outlined in Table 4. Half of patients (48.5%) received glucocorticoids during admission.

**Table 4:** Treatment and outcomes

|  | Items | Total, N (103) |
| --- | --- | --- |
| Treatments | Glucocorticoids, n (%) | 50 (48.5) |
|  | Hydroxychloroquine, n (%) | 82 (79.6) |
|  | Antiviral Kaletra (combination lopinavir/rapinavir) | 54 (52.4) |
|  | Insulin administered on admission, n (%) | 63 (61.2) |
|  | intravenous insulin | 10 (9.7) |
|  | subcutaneous insulin | 53 (51.5) |
| | Units of insulin per 24 hours mean (SD) | 29.6 (±25.6) |
| Outcomes | Hypoglycaemia, n (%) |  |
|  | hypoglycaemia | 5 (4.9) |
|  | hydroxychloroquine | 2 (1.9) |
|  | Outcomes*, n (%) |  |
|  | Required ICU | 29 (28.2) |
|  | Need ventilation | 12 (11.7) |
|  | Need ECMO | 2 (1.9) |
|  | Total death (all in ICU) | 5 (4.9) |

61.2% patients needed insulin during admission and required an average of 29.6 units of insulin per 24 hours. 5 patients had documented hypoglycaemia (defined as blood glucose level < 4 mmol/L) on capillary blood glucose during admission (2 of them were on hydroxychloroquine; 4 on insulin; and one had end stage renal disease).

5 patients died. All of them were in ICU. 4 of them were of Asian origin, 2 males and 3 females. 3 of them were known to have type 2 diabetes and 2 had new diagnosis of prediabetes on admission. 2 of them were obese, one had end stage renal disease with known breast cancer. All of them had pneumonia and 4 of them developed acute respiratory distress syndrome with septic shock. Their average length of stay in hospital was 18 days.

The Survival curve (Figure 1) showed that out of the 5 deaths occurring among the 103 patients, 4 of these were among the group treated by both treatments’ hydroxychloroquine and glucocorticoids (as might be anticipated based on severity of illness). The median time of death was 21 [3 – 39] days.

**Figure 1.**
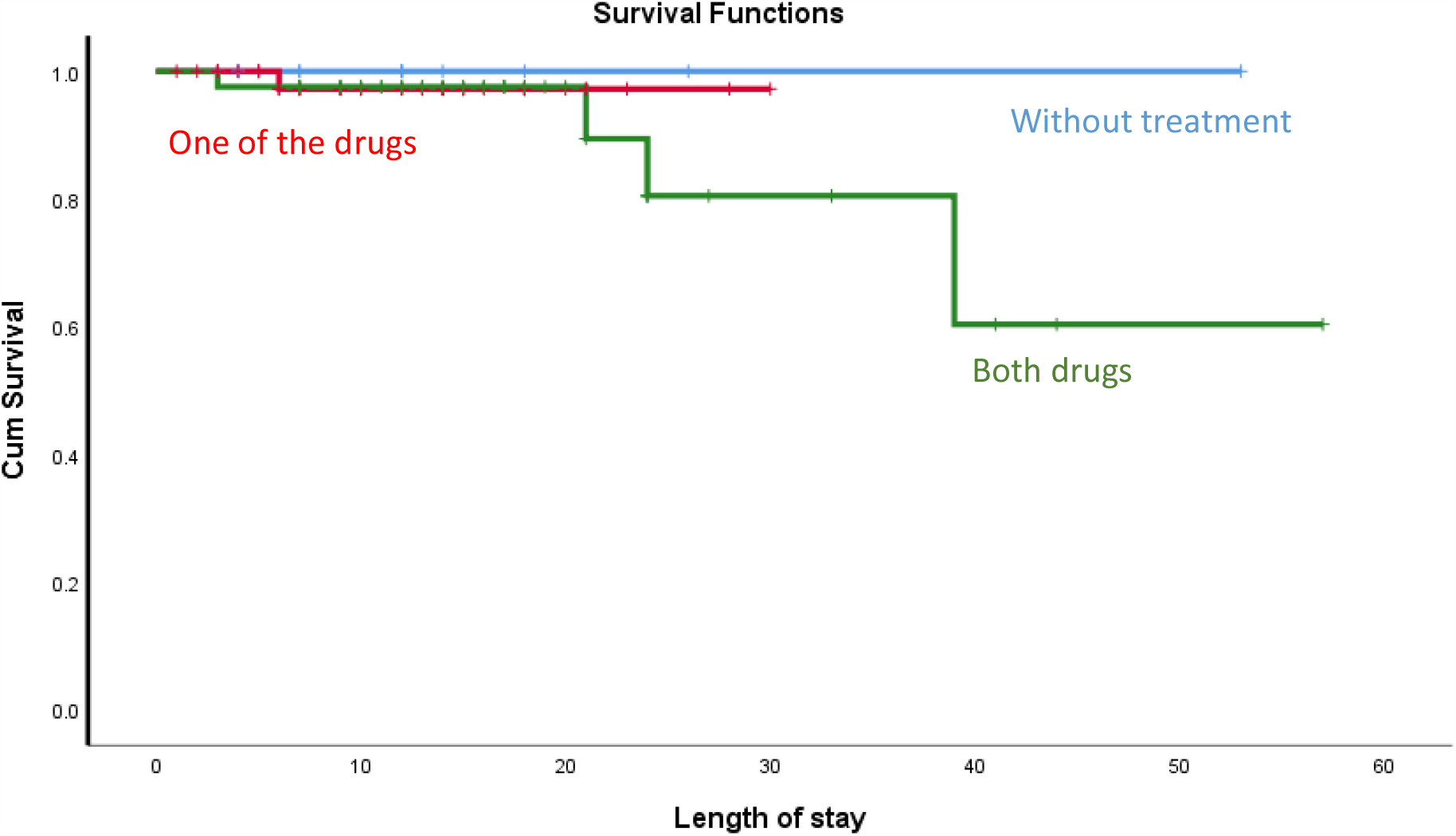
Survival curve using Kaplan Meier Method

### Subset Analysis

We subanalysed the laboratory data for patients on admission and maximum results during their length of stay. Fibrinogen, D-dimer and CRP increased statistically significantly during the course of hospital stay (Table 5).

**Table 5.** Admission and maximal inpatient levels of key biomarkers.

|  | On admission<br>mean (SD) | During admission maximum<br>mean (SD) | P value |
| --- | --- | --- | --- |
| Fibrinogen (mg/dl) | 529.29 (±175.23) | 550.13 (±178.33) | 0.0008 |
| Procalcitonin | 0.63 (±1.24) | 1.50 (±3.20) | 0.12 |
| CRP (mg/L) | 64.15 (±82.7) | 92.2 (±92.45) | 0.000000015 |
| D-dimer (µg/ml) | 1.16 (±2.09) | 2 (±3.98) | 0.01 |
| Ferritin (ng/ml) | 757.34 (±1548.88) | 521 (±1898.92) | 0.06 |

Among 103 patients who were admitted with diabetes/prediabetes and COVID-19, 29 needed ICU care. In this cohort, admission levels of fibrinogen, D-dimer, ferritin and CRP was statistically significantly higher (p<0.05) than in those managed by ward-based care (Table 6).

**Table 6.**
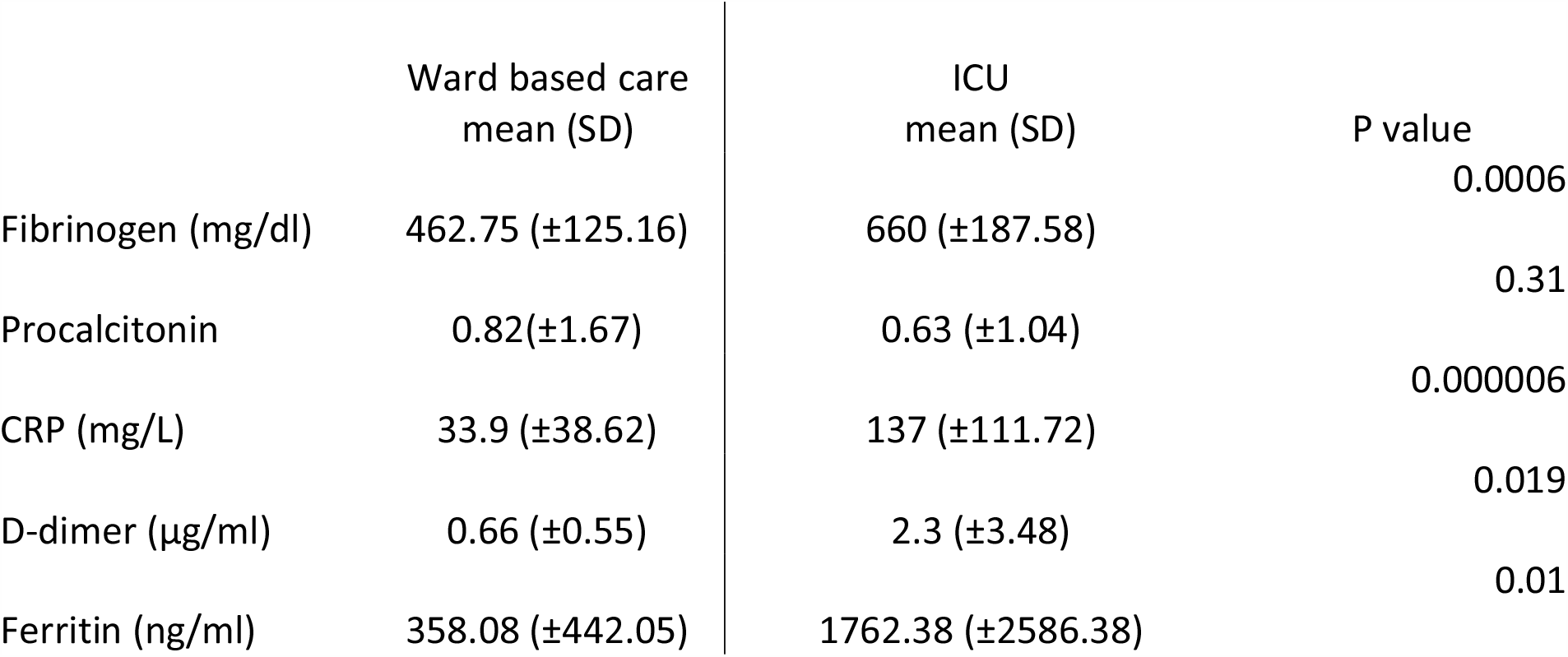
Key biomarkers as markers of severity.

|  | Ward based care<br>mean (SD) | ICU<br>mean (SD) | P value |
| --- | --- | --- | --- |
| Fibrinogen (mg/dl) | 462.75 (±125.16) | 660 (±187.58) | 0.0006 |
| Procalcitonin | 0.82(±1.67) | 0.63 (±1.04) | 0.31 |
| CRP (mg/L) | 33.9 (±38.62) | 137 (±111.72) | 0.000006 |
| D-dimer (µg/ml) | 0.66 (±0.55) | 2.3 (±3.48) | 0.019 |
| Ferritin (ng/ml) | 358.08 (±442.05) | 1762.38 (±2586.38) | 0.01 |

## Discussion

In this single-centre study in Dubai, 25% of patients (n=103) admitted with COVID-19 also had diabetes/prediabetes. 14.6% of this study cohort had new diagnosis diabetes/prediabetes on admission. According to International Diabetes Federation (IDF), prevalence of diabetes in UAE is 17.3% ^8^ and our data reflects this high background prevalence.

Most diabetes patients admitted to hospital with COVID-19 disease were males (67% [n=69]). This agrees with emerging studies showing that men with COVID-19 are at higher risk for developing severe outcomes including death than women. ^3,4,9^ Patients admitted with COVID-19 and diabetes represented 17 different ethnicities (Dubai is a very multi-national society with more than 200 nationalities). Of these, 59.2% (n=61) were Asians and 35% (n=36) were from Arab countries. This finding is also consistent with other studies that suggest that people of Black, Asian, and Minority Ethnic (BAME) populations have increased risk and are predisposed to worser clinical courses and outcomes with COVID-19 infection than Caucasian counterparts.^10^ In addition, many individuals of BAME origin are also more likely to have diabetes/prediabetes. Also, in keeping with recent studies most patients in our cohort with diabetes/prediabetes and COVID-19 infection had other important comorbidities (n=76; 73.8%). This finding is consistent with diabetes and cardiovascular disease being key components of the metabolic syndrome. ^11^ The metabolic syndrome is also associated with pro-inflammatory and pro-thrombotic states which may have important implications for persons affected with COVID-19, where these complications are especially common and troublesome. ^12^

Our laboratory data has shown that fibrinogen, D-dimer, ferritin and CRP levels on admission were higher in those subsequently requiring intensive care treatment. This is reflective of inflammatory cytokine response with COVID-19. Acute hyperglycaemia has shown upregulation of ACE2 gene which facilitates entry of virus inside the cells. With hyperglycaemia over long periods of time it downregulates ACE2 expression making the cells vulnerable to the inflammatory effects of virus.^13^ These laboratory tests can be used on admission to decide the severity of illness and level of care required by the patient (i.e. risk stratification).

Limitations of our study include the small sample size and retrospective nature of analysis. Therefore, more studies with large sample size would help in this guidance. Another limitation of our study is there is no control group of patients without diabetes to compare with (other than knowing absolute numbers of total patients admitted with COVID-19 during the same timeframe). In addition, because of the emergent nature of many new sick patients presenting with COVID-19 to our healthcare facility over a short timeframe, accurate characterisation of overweight/obese status was unavailable for the majority of our patients (and thus are not reported as comorbidities).

## Conclusions

In this single-centre study in Dubai, 25% of patients admitted with COVID-19 also had diabetes/prediabetes. Most diabetes patients admitted to hospital with COVID-19 disease were males of Asian origin. 14.6% had new diagnosis of diabetes/prediabetes on admission. The majority of patients with diabetes/prediabetes and COVID-19 infection had other important comorbidities (n=76; 73.8%). Only 4 patients had negative COVID-19 RT-PCR but had pathognomonic changes of COVID-19 radiologically. Our comprehensive laboratory analysis revealed distinct abnormal patterns of biomarkers that are associated with poor prognosis: Fibrinogen, D-dimer, Ferritin and CRP levels were all statistically significantly higher (p<0.05) at presentation in patients who subsequently needed ICU care compared with those patients who remained ward-based. 28.2% overall needed ICU admission, out of which 5 patients died. More studies with larger sample sizes are needed to compare data of COVID-19 patients admitted with and without diabetes within the UAE region.

## Data Availability

I confirm that all data referred to in this manuscript is available for review.

## Abbreviations

ACE-I: Angiotensin converting enzyme inhibitors
ARB: Angiotensin receptor blocker
ECMO: Extracorporeal membrane oxygenation
HRCT: High resolution computed tomography
RT-PCR: Reverse transcriptase polymerase chain reaction

## References

1. World Health Organization. Coronavirus disease (COVID-19) pandemic [article online]. Available: https://www.who.int [Accessed 3 July 2020].

2. World Health Organization. Coronavirus disease (COVID-2019) situation reports [article online]. Available: https://www.who.int/emergencies/diseases/novel-coronavirus-2019/situation-reports [Accessed 3 July 2020].

3. Zhou F, Yu T,Du R, et al. Clinical course and risk factors for mortality of adult inpatients with COVID 19 in Wuhan, China: a retrospective cohort study. The Lancet. 2020; 395: 1054–62

4. Yang J, Zheng Y, Gou X, Pu K, Chen Z, Guo Q, et al. Prevalence of comorbidities and its effects in patients infected with SARS-CoV-2: a systematic review and meta-analysis. Int J Infect Dis.2020; 94: 91–95.

5. Bornstein SR, Rubino F, Khunti K, et al. Practical recommendations for the management of diabetes in patients with COVID-19. Lancet Diabetes Endocrinol 2020; 8: 546–550.

6. Bode B, Garrett V, Messler J, et al. Glycemic characteristics and clinical outcomes of COVID 19 patients hospitalized in the United States. J Diabetes Sci Technol. 2020; 14: 813–821

7. Barron E, Bakhar C, Kar P, et al. Type 1 and Type 2 diabetes and COVID-19 related mortality in England: a whole population study. https://www.england.nhs.uk/wp-content/uploads/2020/05/valabhji-COVID-19-and-Diabetes-Paper-1.pdf (Accessed July 4th 2020)

8. International Diabetes Federation.https://idf.org/our-network/regions-members/middle-east-and-north-africa/members/49-united-arab-emirates.html (Accessed 3 July 2020)

9. Chen N, Zhou M, Dong X, etal. Epidemiological and clinical characteristics of 99 cases of 2019 novel coronavirus pneumonia in Wuhan, China: a descriptive study. Lancet 2020; 395:507–13.

10. Khunti K, Singh AK, Pareek M, Hanif W. Is ethnicity linked to incidence or outcomes of covid-19? BMJ 2020;369:m1548 doi:10.1136/bmj.m1548

11. Matfin G. The Metabolic syndrome – What’s in a name? Therapeutic Advances in Endocrinology and Metabolism. 2010 1: 39–45

12. Wise J. Covid-19 and thrombosis: what do we know about the risks and treatment? BMJ 2020;369:m2058 doi:10.1136/bmj.m2058

13. Bindom SM, Lazartigues E. The sweeter side of ACE2: physiological evidence of role in diabetes. Mol Cell Endocrinol 2009; 302: 193–202.

